# HepB-Boost: Implementation of free of charge healthcare worker hepatitis B testing and vaccination at Kilifi County Referral Hospital, Kenya

**DOI:** 10.1101/2025.08.27.25334580

**Authors:** Louise Downs, Linus Tinga, Monique Andersson, Philippa C Matthews, Joan Madia, Beatrice Moraa, Liz Gathua, Malik-Ul-Ashtar Tajbhai, Nancy Kagwanja, Nadia Aliyan

**Affiliations:** Nuffield Department of Medicine, University of Oxford, Oxford, UK; KEMRI-Wellcome Trust Research Programme, Kilifi, Kenya; Kilifi County Referral Hospital, Kilifi, Kenya; Oxford University Hospitals NHS Foundation Trust, Oxford, UK; Radcliffe Department of Medicine, University of Oxford, Oxford, UK; Division of Infection and Immunity, University College London, London, UK; The Francis Crick Institute, 1, Midland Road, London, UK; Department of Infectious Diseases, University College London Hospital, Euston Road, London NW1 2BU, UK; Nuffield Department of Primary Care Health Sciences, University of Oxford, Oxford, UK

**Keywords:** Hepatitis B, healthcare worker, vaccination, occupational health, implementation

## Abstract

**Objective:** Healthcare workers (HCWs) are at high risk of HBV exposure due to contact with blood and bodily fluids. In Kenya, HCWs are rarely fully vaccinated against HBV. During 2024, Kilifi County Referral Hospital (KCRH) on the Kenyan coast implemented HBV testing and vaccination for KCRH HCWs.

**Methods:** A technical working group was formed at KCRH to implement and assess a new HBV HCW vaccination programme. Sensitisation was undertaken and vaccines were procured in multi-dose vials. Clinics were run for two hours twice weekly over six-months. HBsAg testing was available, and vaccination offered at 0, 1 and 6 months. Cost analysis was undertaken along with an assessment of vaccination feasibility within existing hospital infrastructure.

**Findings:** 366/574 (64%) of staff received at least one vaccine. Of those attending, the number of fully vaccinated staff increased from 189/366 (5%) to 164/366 (45%), with 289/366 (79%) receiving at least two vaccines. 125/366 (35%) were tested for HBsAg among whom 4/125 (3%) tested positive. Previous HBV vaccination and opting for HBsAg testing were associated with vaccination completion. The estimated cost of the vaccination programme was $4176 with each fully vaccinated person costing $25.45. The primary obstacle to vaccination in this programme was a national shortage of HBV monovalent vaccine in multi-dose vials.

**Conclusions:** HBV vaccination for HCWs was feasible and acceptable at KCRH and could be offered at other similar sized hospitals. Consistent access to HBV monovalent vaccine must be a priority for Kenya, and HCW occupational health screening for HBsAg should be routine.

## Introduction

Healthcare workers (HCWs) in Kenya have a considerable risk of hepatitis B virus (HBV) exposure through contact with blood and bodily fluids. One study at Kenyatta National Hospital, Nairobi reported 14% of staff had sustained a needlestick injury (NSI) in the previous year, and nearly 40% had sustained one throughout their career (1). Despite HBV vaccination being included in Kenyan National Immunisation guidelines since 2007, uptake amongst HCWs remains low. Vaccination is not mandatory, and routine HCW occupational health assessment and vaccine access is inconsistent. Some studies from Kenyan healthcare facilities have reported <50% of their staff are fully vaccinated (2,3).

More attention on HBV prevention is urgently required to support progress towards international elimination targets, and particular investment will be required for the WHO African region (WHO-AFR). Approximately 70% of the world’s new HBV infections occur here, however only 4.2% of those infected with HBV are diagnosed and 0.2% are on treatment (4). In Kenya HBsAg seroprevalence is between 3-8% (5) and can be much higher in HCWs, with few knowing their HBV serostatus (6).

Kilifi County Referral Hospital (KCRH) is on the Kenyan coast, employing approximately 500 HCWs. There is no formal assessment of HCW HBV serostatus or vaccination. We formed an HBV technical working group (TWG) at KCRH consisting of management, clinicians, pharmacists, laboratory staff and nurses, aiming to implement free of charge HBV testing and vaccination for all HCWs employed at KCRH. We describe the design and implementation of this intervention, evaluate its feasibility, and identify lessons for similar programmes in low resource settings.

While many studies in WHO-AFR have assessed HBV vaccination coverage among HCWs, research detailing the processes and outcomes of program implementation is lacking. Such studies could provide valuable insights into overcoming logistical challenges, improving vaccine accessibility, and enhancing awareness and education efforts tailored to healthcare settings. To our knowledge this is the first study in WHO-AFR to plan and implement HCW HBV vaccination in a structured way in a government hospital.

## Methods

Methods for this study were developed using the MRC/NIHR process evaluation framework: https://www.ukri.org/wp-content/uploads/2021/11/MRC-291121-PHSRN-ProcessEvaluationSummaryGuidance.pdf. The study has been reported in line with the Standards for Reporting Implementation Studies (StaRI) checklist (7). The definition of HCW refers to anyone at KCRH who may encounter patients or bodily fluids including clinicians, nurses, pharmacists, laboratory staff and support staff such as cleaners and porters.

### Study Site

KCRH serves a population of around 1.5 million people (8). At the time of implementation, awareness of HBV at KCRH had been heightened due to a concurrent research study testing hospital attendees for HBV (9), and should be considered when evaluating outcomes. Many members of the HBV TWG had been involved in the research study.

### Targeted Sites and Resources

HBV vaccination was aimed at all departments in KCRH (table 1), with vaccines being delivered in MCH clinics where there is cold chain storage, and staff experienced in vaccine administration. HBsAg testing was made available at the hospital laboratory, or at the vaccination clinic prior to vaccination. A custom password protected database domain was purchased from TrueHost (https://truehost.co.ke/) including ID protection to collect information on vaccinees and to enable vaccine reminders to be sent. The database was designed by the TWG, and the hospital data manager set this up. Information collected from each vaccinee included:

- Demographics
- Vaccines previously received, and number required
- Attendance for HBsAg testing and test results
- Employment information (role in KCRH, years worked at KCH, needlestick or splash injuries at work in the last 12 months)
- Completion of recommended vaccination course.

**Table 1:**
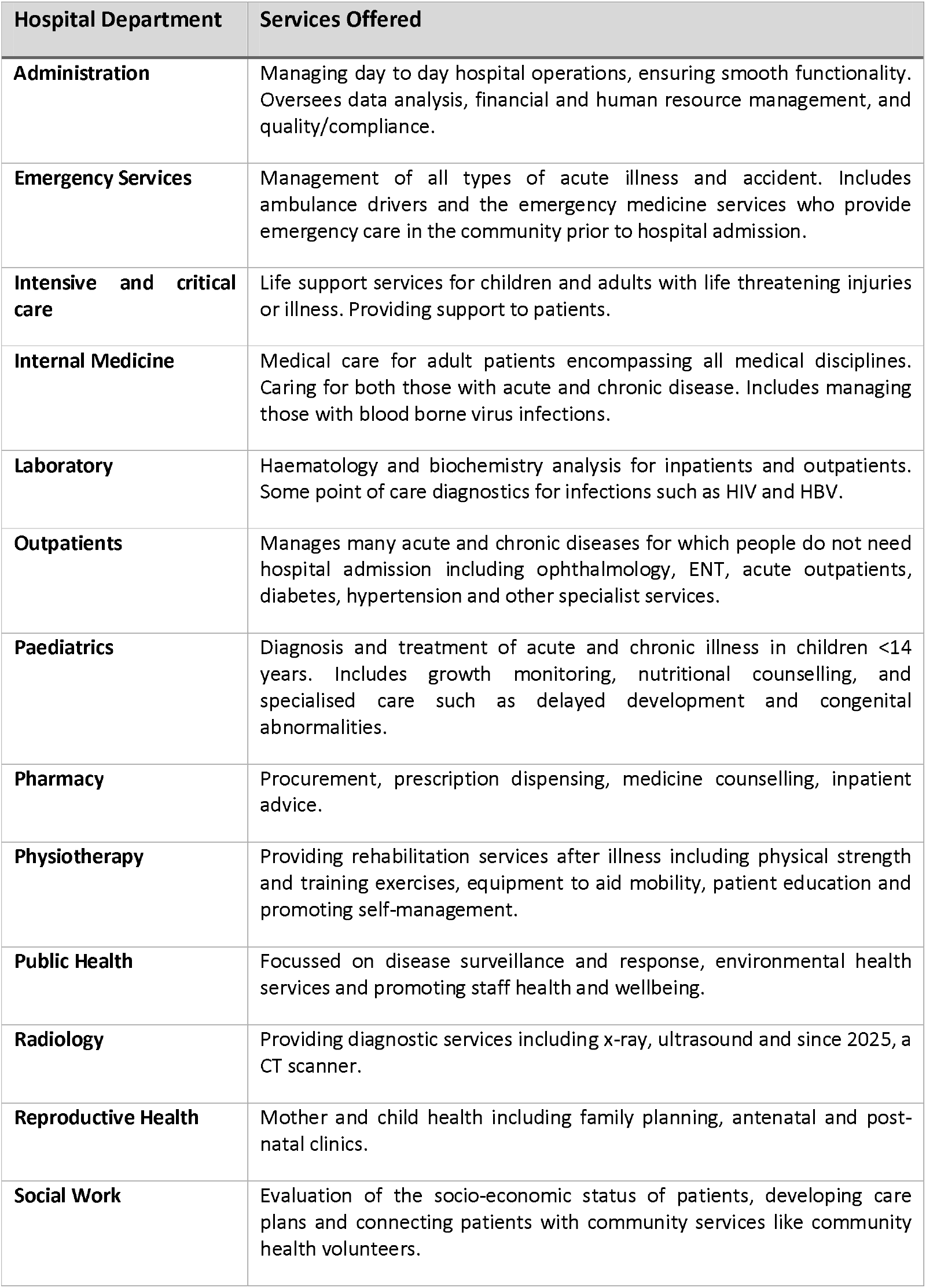
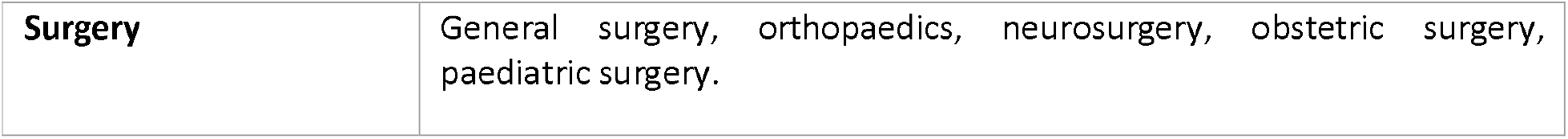
Detailing hospital departments and services provided within each department at Kilifi County Referral Hospital (KCRH), Kenya involved in a drive to provide hepatitis B (HBV) vaccination to healthcare workers (HCWs).

| Hospital Department | Services Offered |
| --- | --- |
| **Administration** | Managing day to day hospital operations, ensuring smooth functionality. Oversees data analysis, financial and human resource management, and quality/compliance. |
| **Emergency Services** | Management of all types of acute illness and accident. Includes ambulance drivers and the emergency medicine services who provide emergency care in the community prior to hospital admission. |
| **Intensive and critical care** | Life support services for children and adults with life threatening injuries or illness. Providing support to patients. |
| **Internal Medicine** | Medical care for adult patients encompassing all medical disciplines. Caring for both those with acute and chronic disease. Includes managing those with blood borne virus infections. |
| **Laboratory** | Haematology and biochemistry analysis for inpatients and outpatients. Some point of care diagnostics for infections such as HIV and HBV. |
| **Outpatients** | Manages many acute and chronic diseases for which people do not need hospital admission including ophthalmology, ENT, acute outpatients, diabetes, hypertension and other specialist services. |
| **Paediatrics** | Diagnosis and treatment of acute and chronic illness in children <14 years. Includes growth monitoring, nutritional counselling, and specialised care such as delayed development and congenital abnormalities. |
| **Pharmacy** | Procurement, prescription dispensing, medicine counselling, inpatient advice. |
| **Physiotherapy** | Providing rehabilitation services after illness including physical strength and training exercises, equipment to aid mobility, patient education and promoting self-management. |
| **Public Health** | Focussed on disease surveillance and response, environmental health services and promoting staff health and wellbeing. |
| **Radiology** | Providing diagnostic services including x-ray, ultrasound and since 2025, a CT scanner. |
| **Reproductive Health** | Mother and child health including family planning, antenatal and post-natal clinics. |
| **Social Work** | Evaluation of the socio-economic status of patients, developing care plans and connecting patients with community services like community health volunteers. |
| **Surgery** | General surgery, orthopaedics, neurosurgery, obstetric surgery, paediatric surgery. |

### Study Design – Implementation Plan

The implementation programme is described in detail at https://doi.org/10.6084/m9.figshare.29877500.v1, but had three main stages: i) HCW sensitisation, development of vaccination guidelines and other promotional materials; ii) implementation of HBV testing and vaccination free of charge and iii) programme evaluation, identifying where processes were altered, challenges faced and how these were addressed.

#### i) Planning and sensitisation

Educational sessions were delivered through continuing medication education (CME) sessions and in individual departments. A standard operating procedure (SOP) was developed covering vaccination procedure, delivery and storage and data collection (10). Posters and leaflets were made (11) and circulated on WhatsApp (Facebook, Inc., 2020).

Printed notices were distributed around KCRH informing staff of the vaccination programme. Self-reported vaccination status was collected by departmental heads to guide procurement, and the rDNA hepatitis B vaccine (Serum Institute of India, Pune, India) was procured privately through Sai Pharmaceuticals (https://saipharm.com/) in 10 dose multi-vials. Monovalent multidose vaccine was not available through the state corporation Kenya Medical Supplies Authority (KEMSA). Data was collected for 353 staff, 269/353 (76%) staff reporting being unvaccinated, 41/353 (12%) reported one previous vaccine, 20/353 (6%) two previous vaccines and 25/353 (7%) were fully vaccinated, giving a total of around 900 vaccines being required to fully vaccinate all those eligible. Initially 600 vaccines were ordered to ensure vaccines did not expire prior to use.

#### ii) Vaccination implementation

The planned vaccine schedule was 0, 1 and 6 months. HCWs were invited to sign up for vaccination on an electronic sheet or manually through departmental managers. Vaccine clinics began on 6^th^ February 2024 and ran for two hours, twice weekly until May 2024, aiming to deliver first and second doses to those who were unvaccinated. Third dose clinics then ran through August 2024. Those who had received one or two previous vaccines could attend any clinic to complete their course. Each vaccine clinic required five staff - two nurses, one administrator, one data manager and one supervisor. Staff were paid 1000 KES (~$7) to run clinics.

Voluntary hepatitis B surface antigen (HBsAg) testing was offered prior to vaccination using a rapid point of care test (POCT) (ACON Laboratories, CA, USA). Those testing HBsAg negative or choosing not to be tested were offered HBV vaccination based on their self-reported vaccine status as per the schedule in figure 1. Those testing HBsAg positive were referred to the HBV clinic for ongoing care.

**Figure 1.**
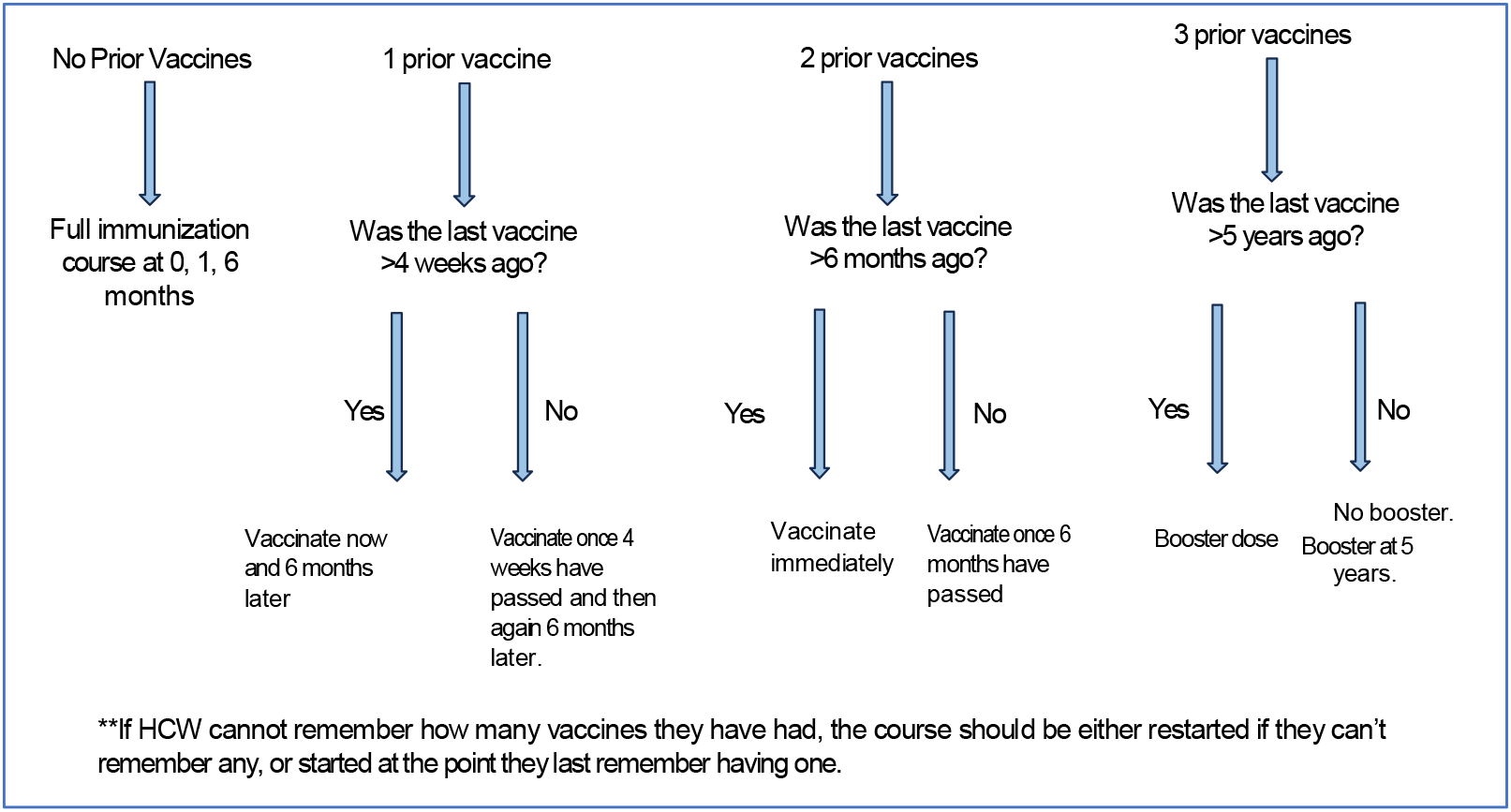
Planned vaccination schedule depending on previous vaccination status at an HBV vaccination drive for staff at Kilifi County Referral Hospital, Kenya. HCW – healthcare worker

Demographics and employment data were collected from vaccinees and entered into the TrueHost database. Future vaccine dates were recorded on the database and physical vaccination cards (12). Text reminders were sent manually prior to the next vaccination dose, avoiding the costs of automatic reminders through a mobile phone provider. Personal phone calls were made if vaccinees did not attend for their scheduled dose.

#### iii) Evaluation

The primary outcome for the HBV vaccination programme evaluation was the number of HCWs tested for and vaccinated against HBV, and factors associated with these outcomes. Further process evaluation objectives are outlined below:

- Did sensitisation reach all target groups?
- Was vaccine procured at a reasonable cost and in a timely manner?
- Was vaccine schedule acceptable and feasible for staff delivering vaccination? Vaccination must not interfere with usual clinic work, and the workload must be manageable for staff delivering vaccine.
- What were the costs of delivering the vaccination programme? Costs were estimated as if vaccines were not donated, and all 366 people who attended for vaccination completed their required schedule, as this is most helpful for planning future programmes.

We aimed to assess the process of implementation by meeting as a TWG every two weeks during the programme.

Multivariable analysis was undertaken to identify individual factors associated with vaccination completion and opting for HBsAg testing. Odds ratios (OR) were calculated, with reference categories being the largest group. For continuous predictor variables, the OR represents the odds of the outcome with each increasing year of age or employment at KCRH. Analysis was undertaken using R version 4.2.0.

We estimated overall programme costs, the cost of one staff member receiving one vaccine, and the cost per fully vaccinated staff member. We also undertook incremental cost effectiveness analysis (ICER) assessing the additional cost incurred to achieve one additional HCW vaccinated. Costs were calculated at the time of implementation in KES, and converted to USD.

Finally, we evaluated the relevance of this implementation in the broader context of providing HBV vaccination to at risk populations in Kenya and further afield.

## Ethics

This implementation programme was undertaken by KCRH in line with existing Kenyan immunisation guidelines (13) so ethical approval was not required. The research initiative alongside this programme to collate and report summary information was approved by the Kenyan Medical Research Institute (KEMRI) Scientific Ethics Review Unit (SERU 4949) and Oxford Tropical Research Ethics Committee (OxTREC 23-24).

## Results

### i) Planning and sensitisation – reaching target groups

One hospital wide CME was undertaken prior to the vaccination programme attended by 40 staff members – mostly clinicians, nurses and pharmacists. Of the 13 attendees who gave feedback, all knew about the HBV vaccine, but 4/13 (30%) did not realise three doses were needed. 12/13 (94%) responders felt the session helped them understand more about the HBV vaccine and all respondents felt the session would encourage vaccination. 10 members of the maternal child health team providing vaccination attended a smaller targeted session, and later in the programme, a tailored session was organised for support staff to encourage attendance.

### ii) Vaccine implementation

Of the estimated 574 people working at KCRH, 366 (64%) received at least one vaccine. Most people attending were women (222/366, 61%) but this may reflect the demographic of HCWs. Age was available for 315/366 people with a median of 34 years (IQR 30-41 years). A summary of vaccinations received by those attending the vaccine clinic is shown in figure 2. The number of staff fully vaccinated against HBV increased from 18/366 (5%) to 164/366 (45%), with 289/366 (79%) of people receiving at least two vaccines (table 2 and figure 2). Anyone due for their 2^nd^ or 3^nd^ vaccine after August 2024 could not complete their required schedule due to lack of vaccine availability.

**Table 2.** Characteristics of those attending hospital vaccination clinics at Kilifi County Referral Hospital as part of a hospital drive to vaccinate employees against HBV infection. Administration includes accountants, IT officers, procurement staff. ^*^Any role containing <10 people was included in ‘Other’, including engineers, drivers, nutritionists, doctors, drivers, social workers, public health officers, security guards, dental officers, physiotherapists and radiographers.

| Characteristic | N | N = 366<sup>1</sup> |
| --- | --- | --- |
| **Age (years)** | 315 | 34 (30, 41) |
| **Sex** | 366 |  |
| Female |  | 222 (61%) |
| Male |  | 144 (39%) |
| **Hospital Role** | 366 |  |
| Accountant |  | 12 (3.3%) |
| Administration |  | 17 (4.6%) |
| Clinical Officer |  | 53 (14%) |
| Laboratory Worker |  | 20 (5.5%) |
| Nurse |  | 141 (39%) |
| Pharmacy |  | 11 (3.0%) |
| Support Staff |  | 72 (20%) |
| Other* |  | 40 (11%) |
| **Needlestick last 12 months** | 366 | 41 (11%) |
| **Years of Service at KCRH** | 366 | 3 (1, 7) |
| **Number of previous vaccines (self reported)** | 366 |  |
| 0 |  | 286 (78%) |
| 1 |  | 42 (11%) |
| 2 |  | 20 (6%) |
| 3 |  | 18 (5%) |
| **Tested for HBV** | 366 |  |
| Yes |  | 125 (34%) |
| No |  | 241 (66%) |
| **HBsAg result** | 125 |  |
| Positive | 4 | (3%) |
| Negative | 121 | (97%) |
| **Completed vaccination schedule** | 366 |  |
| Yes |  | 164 (45%) |
| No |  | 202 (65%) |
| **Vaccination status following the programme** | 366 |  |
| 3 doses (Fully vaccinated) |  | 164 (45%) |
| 2 doses of vaccine received |  | 125 (34%) |
| 1 dose of vaccine received |  | 77 (21%) |

<sup>1</sup>Median (Q1, Q3); n (%)

**Figure 2.**
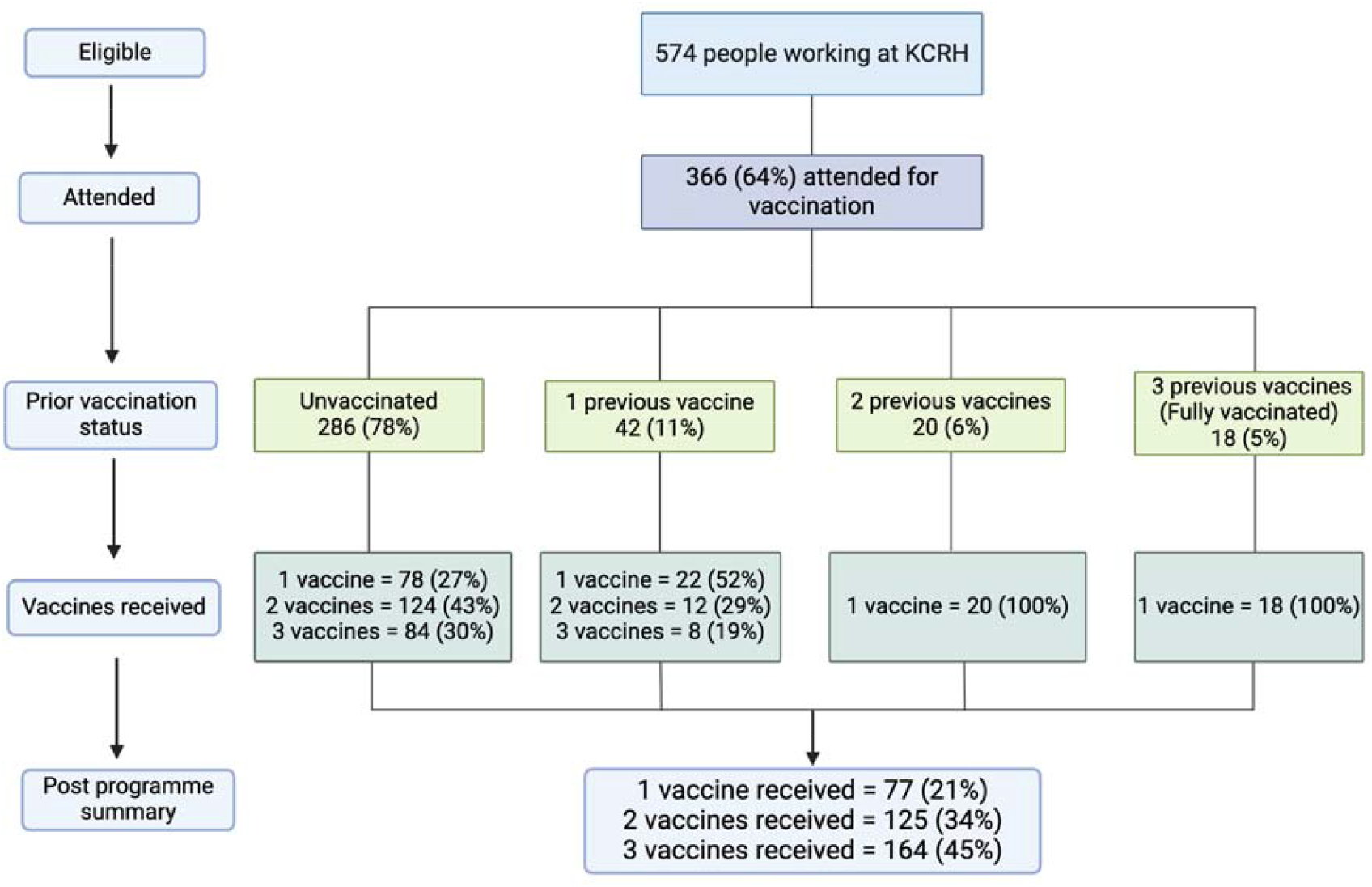
Summarising numbers attending for HBV vaccination, and number of vaccinations received as part of a staff implementation programme at Kilifi County Referral Hospital in Kenya.

Most staff attending for vaccination were nurses (141/366, 39%) or support staff (72/366, 20%). The median length of service at KCRH was 3 years (IQR 1-7 years) and 41 people (11%) reported a needlestick or splash injury in the previous 12 months. 125/366 (34%) of those attending opted for HBsAg testing of whom 4/125 were positive (3.2%) (table 2) and referred for care.

### iii) Multivariable analysis to assess completion of vaccine schedule and HBV testing

There was no association with age, sex or hospital role with completion of vaccination schedule. Those who had at least one previous vaccine and those opting for an HBV test were more likely to complete their vaccination schedule on both univariable and multivariable analysis (table 3).

**Table 3.** Univariable and multivariable analysis of factors associated with completion of hepatitis B vaccination at Kilifi County Referral Hospital as part of a hospital drive to vaccinate staff. Rows in bold indicate significant associations. Reference groups are shown in brackets.

| Predictor (reference group) | UNIVARIABLE ANALYSIS |  |  |  | MULTIVARIABLE ANALYSIS |  |  |  |
| --- | --- | --- | --- | --- | --- | --- | --- | --- |
|  | Odds Ratio | 95% CI Lower | 95% CI Upper | P-value | Odds Ratio | 95% CI Lower | 95% CI Upper | P-value |
| Sex (female) | 0.93 | -0.49 | 0.35 | 0.74 |  |  |  |  |
| Age | 1.02 | -0.01 | 0.04 | 0.18 |  |  |  |  |
| Hospital role (nurse) | 0.75 | -1.55 | 0.89 | 0.63 |  |  |  |  |
| Years of service | 0.99 | -0.04 | 0.02 | 0.58 |  |  |  |  |
| Previous needlestick (no) | 1.20 | -0.48 | 0.83 | 0.59 |  |  |  |  |
| **Previous Vaccine (none)** | **97.00** | **3.06** | **7.46** | **<0.01** | **93.02** | **20.15** | **1655.7** | **<0.01** |
| **HBV test (no)** | **2.20** | **0.35** | **1.24** | **<0.01** | **1.91** | **1.12** | **3.27** | **0.02** |

When investigating factors associated with being tested for HBV, on univariable analysis, age, years of service at KCRH and having been previously vaccinated were significantly associated with having a test, however only a history of vaccination was significant on multivariable analysis (table 4).

**Table 4.**
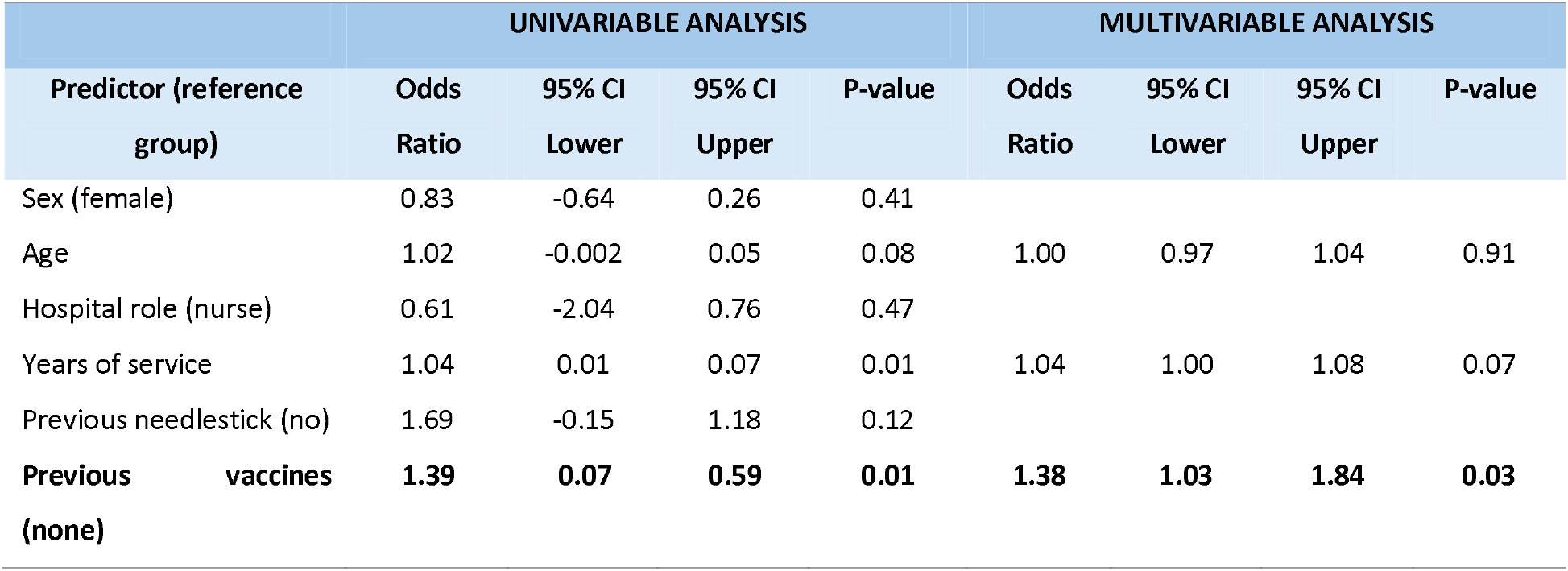
Univariable and multivariable analysis of factors associated with being tested for hepatitis B at Kilifi County Referral Hospital as part of a hospital drive to test and vaccinate staff. Rows in bold indicate significant associations. Reference groups are shown in brackets.

### iv) Vaccine access and delivery feasibility

Following completion of second-dose clinics (in July 2024), vaccine supplies ran out and we were unable to procure more multi-dose vials. Single dose vials were prohibitively expensive (492 KES ($3.80)/dose as opposed to 223 KES ($1.70)/dose). A donation of 200 near-expiry vaccines from KWTRP allowed programme continuation through August 2024, where those still unvaccinated were encouraged to attend, along with those who had presented late and required 2^nd^ and 3^nd^ doses. The donated vaccines expired on 31^st^ August 2024, so at that point the vaccination programme stopped.

Clinics were labour intensive especially in the first few weeks when many people attended and were frequently opened outside of scheduled hours. Significant time was dedicated to following up those who missed their appointments. Use of automatic mobile phone reminders could have reduced this burden and ensured accuracy despite the added cost. Vaccine clinics did not interfere with the day to day running of the MCH department.

### v) Estimated costs of vaccination programme

The estimated costs for this programme are shown in table 5. Multi-dose vaccines were procured at a cost of 2237 KES for 10 doses (~$17 for 10, $1.70/dose). Vaccine equipment was costed along with HBsAg testing kits and airtime for follow up calls and text messages. Infant vaccination clinics already had cold storage, so no new equipment was required. Staff running clinics were reimbursed 1000 KES each (~ $7.70) per clinic (other than the supervisor).

**Table 5.** Estimated costs for delivery of staff hepatitis B vaccination at Kilifi County Hospital, Kenya. Costs are given in both Kenyan Shillings (KES) and US dollars (USD). IT: Information technology; HBsAg – hepatitis B surface antigen.

| Cost Group |  | Specifics | Cost for<br>1 | Total<br>needed | Total Cost<br>(KES) | Total Cost<br>(USD) | Comments |
| --- | --- | --- | --- | --- | --- | --- | --- |
| STAFF |  | Nurse | 1000 | 38 | 76000 | 590 | 2 x vaccine clinics per week for 15 weeks for dose 1 & 2, then 2 clinics per week for 4 weeks for dose 3. |
|  |  | Nurse | 1000 |  |  |  |  |
|  |  | Data manager | 1000 | 38 | 38000 | 294 |  |
|  |  | Administrative staff | 1000 | 38 | 38000 | 294 |  |
|  |  | Supervisor |  | 38 | Nil | Nil | No extra pay |
| IT |  | Database setup and maintenance | 10000 | 1 | 100000 | 774 |  |
|  |  | Database (TrueHost) | 5104 | 1 | 15000 | 116 | Highest level of security |
| CONSUMABLES | Vaccine Promotion | Printing posters | 30 | 20 | 600 | 5 |  |
|  |  | Printing leaflets | 30 | 300 | 9000 | 70 |  |
|  |  | Printing vaccine cards | 15 | 250 | 3750 | 29 |  |
|  | Vaccinations | Sai Pharmaceuticals (10 dose vials) | 2237 | 100 | 223700 | 1730 | Assuming 1000 vaccines required |
|  |  | Needles | 6.54 | 1000 | 6540 | 50 |  |
|  |  | Syringes | 6.94 | 1000 | 6940 | 54 |  |
|  |  | Cotton wool roll | 215 | 4 | 860 | 7 |  |
|  |  | Surgical spirit per ml | 0.18 | 15000 | 2700 | 21 |  |
|  |  | Sharps bin | 95 | 4 | 380 | 3 |  |
|  | HBsAg tests | Medix East Africa Ltd | 32 | 500 | 16000 | 124 |  |
| OTHERS |  | Airtime for follow up (calls + text messages) |  |  | 2000 | 15 |  |
|  |  | Follow up phone calls |  | 8 | No extra | Nil | 2 hours/week for 4 weeks by 2 staff. |
| TOTAL |  |  |  |  | 539470 | 4176 |  |

The total cost of this vaccination programme to deliver 1000 vaccines to staff was 539,470 KES (~$4176). The greatest cost was the vaccinations (223,700 KES, ~$1730, 51%)) followed by staff delivering vaccinations (152,000 KES, ~$1200, 35%). The cost per staff member receiving at least one vaccine was $11.40, whilst the cost per fully vaccinated staff number was $25.45. When looking at the ICER, 146 additional staff members were fully vaccinated after the programme, giving an incremental cost per additional fully vaccinated staff member of $28.59.

### vi) Fidelity and Adaptation

Some alterations were made to the original implementation plan:

- Non patient facing staff were vaccinated. We opted not to turn anyone away for fear of negative repercussions on the programme, however this meant more vaccines were used than anticipated.
- Several staff from peripheral hospitals attended for vaccination. Staff did not have official identification badges at the time of this programme.
- Those who were fully vaccinated many years ago wanted to restart an HBV vaccine course. To reinforce WHO vaccination schedule, we implemented a clinical supervisor at each clinic to ensure those incompletely vaccinated were prioritised.

We would suggest in future programmes, vaccination provision should be stratified depending on risk, and history of vaccination to ensure those in greatest need are prioritised.

### vii) Contextual Changes

During this implementation there was a change in hospital management meaning repeat sensitisation, and differing priorities for the use of hospital resources. Along with this, HBV multi-dose monovalent vaccine became nationally unavailable over the course of the study. Despite donations from KWTRP, this limited number of staff who could be completely vaccinated.

## Discussion

We have delivered an HBV vaccine programme at KCRH, and evaluated the process, feasibility and cost of implementation. There was excellent engagement from the TWG and maternal child health teams throughout despite meetings and clinics being intensive in the first few weeks.

Of the 64% of KCRH staff who attended a vaccination clinic, 76% were unvaccinated prior to this programme, and following this implementation, 79% had received at least two vaccines. Those attending were a self-selecting population and staff who did not attend may have been already fully vaccinated or disengaged with healthcare and not vaccinated at all. Studies from other hospitals in Kenya have reported varying vaccination levels. One study covering a sub selection of HCWs from eight county hospitals around Nairobi showed 80% of staff had received at least one vaccine, and 48% were fully vaccinated (3), whilst another study assessing all HCWs from two hospitals in Thika District in Central Kenya showed 87% HCWs had never been previously vaccinated (14).

Sensitisation efforts during the vaccination programme reached many hospital groups; however, support staff were initially underrepresented due to limited internet access. These groups are at high risk of NSI as they handle hospital waste, which may contain improperly discarded sharps.

Targeted sensitisation sessions increased attendance later in the programme, however these strategies should have been employed from the outset, organised independently of digital messaging.

Those who had received at least one previous HBV vaccine and those opting for HBsAg testing were significantly more likely to complete their vaccination schedule, and those previously vaccinated were more likely to opt for HBsAg testing than those unvaccinated. These people are likely to be more engaged in healthcare and have a greater understanding of the need for testing and vaccination.

Only 34% of those attending for vaccination accepted free HBsAg testing. We intentionally made this optional to avoid deterring individuals from seeking vaccination. While testing was mentioned during sensitisation sessions, the primary focus was vaccination benefits. Moving forward, we recommend placing greater emphasis on the advantages of HBsAg testing and suggest having a clinical advisor available dedicated to encouraging testing.

The most significant barrier to vaccination schedule completion was unavailability of multi-dose vials. Single dose vials were over twice the price per dose and therefore not feasible. Cuts to global health funding by international governments makes future vaccine access even more uncertain. The US administration recently terminated a $2.6 billion grant to the Global Vaccine Alliance (GAVI) (15) which will severely impact the availability of vaccines to low and middle income countries like Kenya. Steps must be made by vaccine suppliers to ensure equitable, affordable access, whilst efforts to fill funding gaps are consolidated.

Whilst this programme was labour intensive, direct costs were relatively small and represent outgoings for initial set up. Future costs would be lower as most staff would already be vaccinated, and pathways would already be established. The cost metrics presented are to aid preliminary budget planning and felt to be most useful for Kenyan hospitals currently. Moving forwards a more detailed cost effectiveness analysis would consider direct costs avoided for acute HBV treatment, along with savings later in life due to sequelae from chronic HBV infection. One study from Ethiopia modelled the costs of increasing HBV HCW vaccination from 14% (as is the current situation) to 80% as recommended by WHO (16), showing mandatory vaccination was cost effective. Another study from Ethiopia found that the majority of HCWs were willing to pay for HBV vaccination, however less than the market price, suggesting that healthcare facilities could improve access by offering vaccines at subsidised cost rather than covering the full expense (17).

The end goal of programmes such as this is provision of routine hospital occupational health services, involving blood borne virus testing and review of vaccination history. Allocation of an occupational health specialist may not be possible for many Kenyan County hospitals but should be considered in future health budget planning.

## Conclusions and Implications

Implementing HBV testing and vaccination for HCWs in KCRH using existing infrastructure was feasible, and is scalable to other hospitals, however, requires ongoing sensitisation and a dedicated team. Ensuring consistent access to affordable HBV vaccines must be a priority WHO-AFR to enable protection of at-risk groups and make headway towards global elimination targets.

## Data Availability

For the purpose of Open Access, the author has applied a CC-BY public copyright license to any author accepted manuscript version arising from this submission. Data supporting the findings of this study will be publicly available on the acceptance of the manuscript for publication. This manuscript was written with the permission of Director KEMRI CGMRC.

## Acknowledgements

We would like to thank the entire vaccination team at Kilifi County Hospital for their enthusiasm and dedication to this programme, and special mention should go to Linah Mwasho, Customer Care at KCRH. Without you all, this would not have been possible.

